# Triglyceride/High Density Lipoprotein Cholesterol Index and Future Cardiovascular Events in Diabetic Patients without Known Cardiovascular Disease

**DOI:** 10.1101/2024.08.30.24312870

**Authors:** Ryosuke Nakashima, Shota Ikeda, Keisuke Shinohara, Sho Matsumoto, Daisuke Yoshida, Yoshiyasu Ono, Hiroka Nakashima, Ryohei Miyamoto, Shouji Matsushima, Junji Kishimoto, Hiroshi Itoh, Issei Komuro, Hiroyuki Tsutsui, Shintaro Kinugawa, Kohtaro Abe

**Affiliations:** Department of Cardiovascular Medicine, Faculty of Medical Sciences, Kyushu University, Fukuoka, Japan; Center for Clinical and Translational Research, Kyushu University Hospital, Fukuoka, Japan; Center for Preventive Medicine, School of Medicine, Keio University, Tokyo, Japan; Department of Frontier Cardiovascular Science, The University of Tokyo Graduate School of Medicine, Tokyo, Japan; International University of Health and Welfare, Tokyo, Japan; International University of Health and Welfare, Fukuoka, Japan

**Keywords:** Triglyceride, High-density lipoprotein cholesterol, Diabetes, Dyslipidemia, Statin

## Abstract

**Background:** The triglyceride/high-density lipoprotein cholesterol (TG/HDL-C) index, calculated as TG levels divided by HDL-C levels, is suggested as a predictor of cardiovascular disease (CVD). We investigated the association between the TG/HDL-C index and initial CVD events in high-risk patients with type 2 diabetes mellitus (T2DM).

**Methods:** We analyzed the association between the TG/HDL-C index and CVD events in T2DM patients with retinopathy and hyperlipidemia without known CVD enrolled in the EMPATHY study, which compared intensive and standard statin therapy (targeting LDL-C levels <70 mg/dL and ≥100 to <120 mg/dL, respectively).

**Results:** A total of 4665 patients were divided into high (TG/HDL-C ≥2.5, *n*=2013) and low (TG/HDL-C <2.5, *n*=2652) TG/HDL-C index groups. During a median follow-up of 36.8 months, 260 CVD events occurred. The high TG/HDL-C index group had higher CVD risk compared to the low TG/HDL-C index group (HR 1.89, 95% CI 1.45–2.47, *p*<0.001). This association was consistent across various subgroups. However, a trend toward interaction between the TG/HDL-C index and EMPATHY treatment allocation for CVD risk was observed (*p* for interaction=0.062). Intensive statin treatment was associated with reduced CVD risk compared to standard treatment in the high TG/HDL-C index group but not in the low group.

**Conclusions:** A TG/HDL-C index ≥2.5 was associated with increased CVD risk in T2DM patients with retinopathy and hyperlipidemia without a history of CVD. The TG/HDL-C index may be a predictor of CVD and indicate patients who could benefit from intensive statin treatment.

## Introduction

Type 2 diabetes mellitus (T2DM) is a crucial risk factor for cardiovascular disease (CVD), with CVD being the leading cause of mortality in patients with T2DM.^1,2^ Dyslipidemia is another significant risk factor for CVD. Importantly, the coexistence of T2DM and dyslipidemia substantially increases the risk of CVD. Therefore, risk assessment, prevention, and early detection of CVD are essential in the management of patients with T2DM and dyslipidemia.^3,4^

Among the blood lipid components, high levels of low-density lipoprotein cholesterol (LDL-C) are well recognized as a risk for CVD and a therapeutic target. While strict LDL-C lowering therapy such as intensive statin therapy is well established, attention has focused on the management of lipid components other than LDL-C to reduce the residual risk of CVD. Both high levels of triglyceride (TG) and low levels of high-density lipoprotein cholesterol (HDL-C) are considered risk-enhancing factors for CVD.^1^ High levels of TG, particularly fasting TG levels of ≥150 mg/dL, are associated with an increased risk of CVD.^1,5^ Notably, lipid-lowering therapy aimed at lowering TG, such as fibrate therapy, has shown limited efficacy in the setting of hypertriglyceridemia.^6–8^ Taken together with the fact that, unlike LDL-C, TG does not accumulate in atherosclerotic plaques in the vessel wall,^9^ it is reasonable to assume that TG itself does not promote atherosclerosis, but that the condition causing lipid metabolism disorder contributes to the development of atherosclerosis and CVD. Low levels of HDL-C, particularly <40 mg/dL in men and <50 mg/dL in women, have also been suggested to be associated with increased CVD risk; however, the evidence linking HDL to increased CVD risk is not as robust as the evidence linking TG to increased CVD risk, and there are even reports suggesting that higher-than-normal levels of HDL may increase CVD risk.^1,10^ Given this background, it is desirable to establish useful indices to reduce the residual risk of CVD in dyslipidemia.

Recently, the TG/HDL-C index, which is calculated as TG levels divided by HDL-C levels, has attracted attention as a more sensitive marker for predicting the risk of developing CVD.^11–13^ In a limited population with high suspicion of coronary artery disease undergoing coronary angiography, the association of TG/HDL-C index ≥2.5 with a high risk of CVD was demonstrated.^14^ However, the significance of the TG/HDL-C index in assessing CVD risk in a larger high-risk population, particularly patients with T2DM and hyperlipidemia but without known CVD, is unclear.

The aim of this study was to investigate whether the TG/HDL-C index is associated with CVD events in high-risk T2DM patients with diabetic microvascular complication(s) without known CVD under statin treatment. We used the dataset from the EMPATHY trial, which compared intensive and standard statin treatment in T2DM patients with diabetic retinopathy and hyperlipidemia without a history of CVD.^15^ While the original EMPATHY trial failed to demonstrate a reduction in the risk of CVD events with intensive versus standard statin treatment in this T2DM population,^15^ we performed an additional exploratory analysis to evaluate the effectiveness of intensive versus standard statin treatment in patients stratified by the TG/HDL-C index.

## Methods

### Patients

The subjects of this study are patients enrolled in the EMPATHY study (Clinical Trial Registry: URL, https://www.umin.ac.jp/ctr.; unique identifier, UMIN000003486).^15^ The study was conducted in accordance with the Declaration of Helsinki and the Japanese ethical guidelines for clinical research, and was approved by the ethical committee of Kyushu University (No. 2020-445). Patients with T2DM who had diabetic retinopathy and elevated LDL-C levels (>120 mg/dL without lipid-lowering medication or >100 mg/dL with lipid-lowering medication) were enrolled in the EMPATHY study (N=5,042). Patients with prior coronary artery disease or stroke, symptomatic heart failure (New York Heart Association class IIM or higher), familial hypercholesterolemia, and renal failure (serum creatinine ≥ 2.0 mg/dL or estimated glomerular filtration rate [eGFR] <30 mL/min/1.73 m^2^) were excluded. During the run-in period (4–8 weeks), patients received statin monotherapy with a target of LDL-C ≥100 to <120 mg/dL, and were then randomly assigned to standard statin therapy targeting LDL-C ≥100 to <120 mg/dL or enhanced statin therapy targeting LDL-C <70 mg/dL. In this study, patients were excluded if they did not have values of TG, HDL-C, LDL-C, hemoglobin A1c (HbA1c), eGFR, and brain natriuretic peptide (BNP) at the time of EMPATHY treatment allocation.

### Blood test and TG/HDL-C index

Blood samples were collected under fasting conditions. Blood test measurements, including TG, HDL-C, LDL-C, serum creatinine, HbA1c, and BNP levels, were performed at a central laboratory (SRL, Inc., Tokyo, Japan). The values of TG and HDL-C at the time of EMPATHY treatment allocation were used in this study. The TG/HDL-C index was calculated as the TG level divided by the HDL-C level.

### Outcome

CVD events in this analysis were defined as the composite incidence of cardiovascular events or death associated with cardiovascular events, including coronary, cerebral, renal, and vascular events, which was the same as the primary outcome of the EMPATHY trial.^15^ Coronary events included myocardial infarction, unplanned hospitalization for unstable angina, and coronary revascularization. Cerebral events included ischemic stroke and cerebral revascularization. Renal events included initiation of chronic dialysis or increase in serum creatinine level by at least twofold and ≥1.5 mg/dL from the value at enrollment. Vascular events were defined as aortic or peripheral arterial disease consisting of aortic dissection, mesenteric artery thrombosis, severe lower extremity ischemia (ulcer), revascularization, or finger or lower extremity amputation due to arteriosclerosis obliterans. In this study, we additionally performed analyses for the non-renal cardiovascular events (composite of cardiac, cerebral and vascular events or death associated with these events) and renal events.

### Statistical analysis

Continuous variables were expressed as means and standard deviations (SDs) and analyzed using the t-test, unless otherwise noted. All categorical variables were expressed as raw numbers and percentages and were compared using the chi-square test.

The subjects were divided into two groups; TG/HDL-C index ≥2.5 and TG/HDL-C index <2.5 based on previous literatures.^14^ The Kaplan–Meier cumulative incidence curves of the two groups were compared using the log-rank test. Univariate and multivariate Cox proportional hazards models were used to calculate the hazard ratio (HR) for the incidence of events. We constructed 4 models to adjust for confounding factors. Model 1 included sex, age, and EMPATHY allocation. Model 2 additionally included body mass index (BMI), smoking status, hypertension, systolic blood pressure, LDL-C, HbA1c, and eGFR. Model 3 included BNP (logarithm conversion), diabetic neuropathy or/and diabetic nephropathy. Model 4 included the types of medications: angiotensin II receptor blocker (ARB) or angiotensin converting enzyme inhibitor (ACEI), calcium channel blocker (CCB), diuretic, and β-blocker. In the multivariate analyses examining the association between the TG/HDL-C index and CVD events, we used not only the categorical variables of the TG/HDL-C index with a cutoff of 2.5, but also its continuous variables. To elucidate the association between the TG/HDL-C index and the level of TG and/or HDL-C, we further performed multivariate Cox regression analysis with a model including the covariates included in Model 4 and TG and/or HDL-C. P < 0.05 was considered statistically significant.

Subgroup analysis was performed according to sex, age ≥/<65 years, BMI ≥/<25 kg/m^2^, systolic blood pressure ≥/<140 mmHg, eGFR ≥/<60 mL/min/1.73 m^2^, HbA1c ≥/<7.0%, TG ≥/<118 mg/dL, and HDL-C ≥/<54 mg/dL. The cutoff values for TG and HDL-C were median values in order to examine the influence of the absolute values of the variables rather than the ratios represented by the TG/HDL-C index. Forest plots were generated with high and low TG/HDL-C index. In additional exploratory analyses, Kaplan-Meier curves of CVD events by EMPATHY allocation were plotted according to high and low TG/HDL-C index, and multivariate Cox regression analysis was also performed using a model excluding TG/HDL-C index category from Model 4.

All analyses were conducted with SAS 9.4 (SAS Institute Inc., Cary, NC, USA).

## Results

### Study participants and patient characteristics according to the TG/HDL-C index

The EMPATHY study included 5042 patients, and 377 patients were excluded due to the lack of sufficient data. A total of 4665 patients were analyzed in this study. All CVD events (n = 260) were observed during the median follow-up period of 36.8 (range, 0.03–65.7) months. **Table 1** shows the baseline demographic and clinical characteristics of patients in high (TG/HDL-C index ≥2.5) and low (TG/HDL-C index <2.5) TG/HDL-C index groups. The high TG/HDL-C index and low TG/HDL-C index groups included 2013 and 2652 patients, respectively. Patients in the high TG/HDL-C index group were younger, had higher proportion of male sex, had higher blood pressure, and higher smoking rates. Patients in the high TG/HDL-C index group also had higher TG and lower HDL-C levels. There were no significant differences in LDL-C levels and in EMPATHY treatment allocation between the two groups. ARB, ACEI, diuretic, CCB, and β-blocker were more frequently used in patients with a high TG/HDL-C index.

**Table 1.** Baseline characteristics in the high and low TG/HDL-C index groups.

| | TG/HDL-C index $\geq 2.5$<br>n = 2013 | TG/HDL-C index $< 2.5$<br>n = 2652 | P value |
| --- | --- | --- | --- |
| Age, y | 61.9 ± 11.1 | 63.8 ± 10.1 | < 0.001 |
| Male, n (%) | 1052 (52.3) | 1182 (44.6) | < 0.001 |
| BMI, kg/m<sup>2</sup> | 26.6 ± 4.2 | 25.0 ± 4.3 | < 0.001 |
| Systolic BP, mmHg | 135.6 ± 16.9 | 134.1 ± 16.3 | 0.002 |
| Diastolic BP, mmHg | 75.8 ± 11.6 | 74.1 ± 11.1 | < 0.001 |
| Current smoking, n (%) | 468 (23.3) | 399 (15.1) | < 0.001 |
| Hypertension, n (%) | 1490 (74.0) | 1811 (68.3) | < 0.001 |
| Duration of T2DM, y | 12.5 ± 8.5 | 13.4 ± 9.0 | 0.001 |
| eGFR, mL/min/1.73 m<sup>2</sup> | 72.4 ± 21.2 | 75.4 ± 19.6 | < 0.001 |
| HbA1c, % | 5.6 ± 0.6 | 5.6 ± 0.6 | < 0.001 |
| LDL-C, mg/dL | 113.3 ± 26.1 | 106.5 ± 24.8 | 0.68 |
| HDL-C, mg/dL | 47.1 ± 9.3 | 62.2 ± 13.3 | < 0.001 |
| TG, mg/dL | 203.5 ± 97.0 | 91.3 ± 27.6 | < 0.001 |
| BNP, pg/dL* | 13.1 (6.7, 27.3) | 16.2 (8.3, 30.6) | 0.014 |
| ACEI or ARB | 1111 (55.2) | 1311 (49.4) | < 0.001 |
| CCB | 797 (39.6) | 912 (34.4) | < 0.001 |
| β-blocker | 115 (5.7) | 107 (4.0) | 0.008 |
| Diuretic | 263 (13.1) | 289 (10.9) | 0.023 |
| MRA | 31 (1.5) | 21 (0.8) | < 0.001 |

Values are numbers (%) or mean ± SD except for BNP. \*Median value and interquartile range are shown in parentheses. ACEI, angiotensin-converting enzyme inhibitor; ARB, angiotensin II receptor blocker; BMI, body mass index; BNP, B-type natriuretic peptide; BP, blood pressure; CCB, calcium channel blocker; eGFR, estimated glomerular filtration rate; HbA1c, hemoglobin A1c; HDL-C, high-density lipoprotein cholesterol; LDL-C, low-density lipoprotein cholesterol; MRA, mineralocorticoid receptor antagonist; T2DM, type 2 diabetes mellitus; TG, triglyceride.

### Comparison of the incidence of CVD events between the high and low TG/HDL-C index groups

**Figure 1** shows the Kaplan-Meier curves comparing the cumulative incidence of CVD events between the high and low TG/HDL-C index groups. The incidence of CVD events was significantly higher in the high TG/HDL-C group than in the low TG/HDL-C index group (P < 0.001, log-rank test).

**Figure 1.**
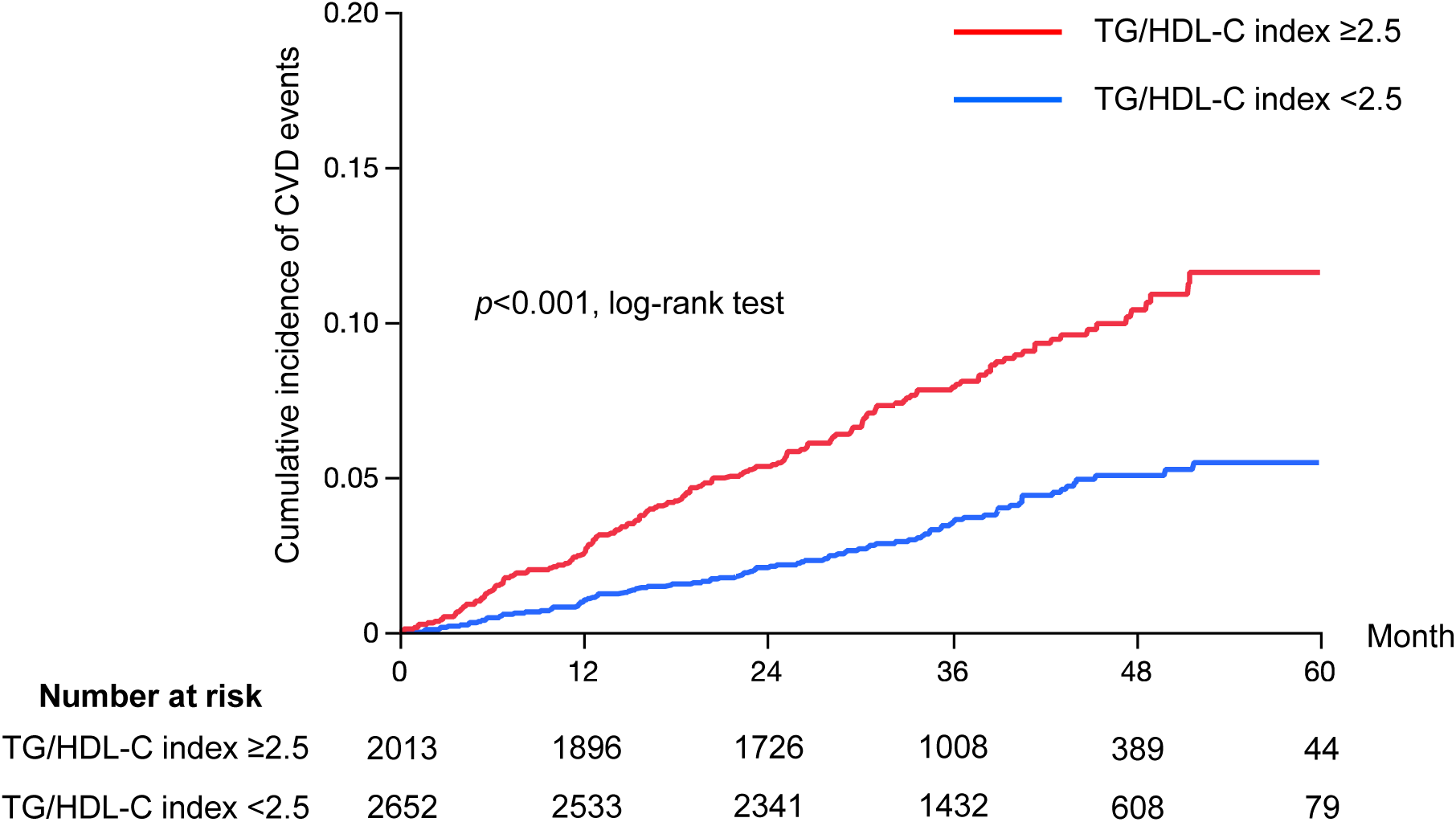
Cumulative incidence of CVD events in the high and low TG/HDL-C index groups. The Kaplan-Meier curves comparing the cumulative incidence of CVD events between the high and low TG/HDL-C index groups. CVD, cardiovascular disease; HDL-C, high-density lipoprotein cholesterol; TG, triglyceride.

In univariate analysis **(Table 2)**, high TG/HDL-C index was associated with the risk of CVD events (HR 2.29, 95% confidence interval [CI] 1.78–2.94, P < 0.001, number of events: TG/HDL-C index <2.5, 98/2652; TG/HDL-C index ≥2.5, 162/2013). Multivariate Cox regression analysis **(Table 2)** showed that a high TG/HDL-C index was associated with a higher incidence of CVD events irrespective of the multivariate models (HR 1.88, 95% CI 1.44-2.45, P < 0.001 in Model 4). This significant association was also observed in the analyses that dealt with the continuous variable as well as with the categorical variable.

**Table 2.**
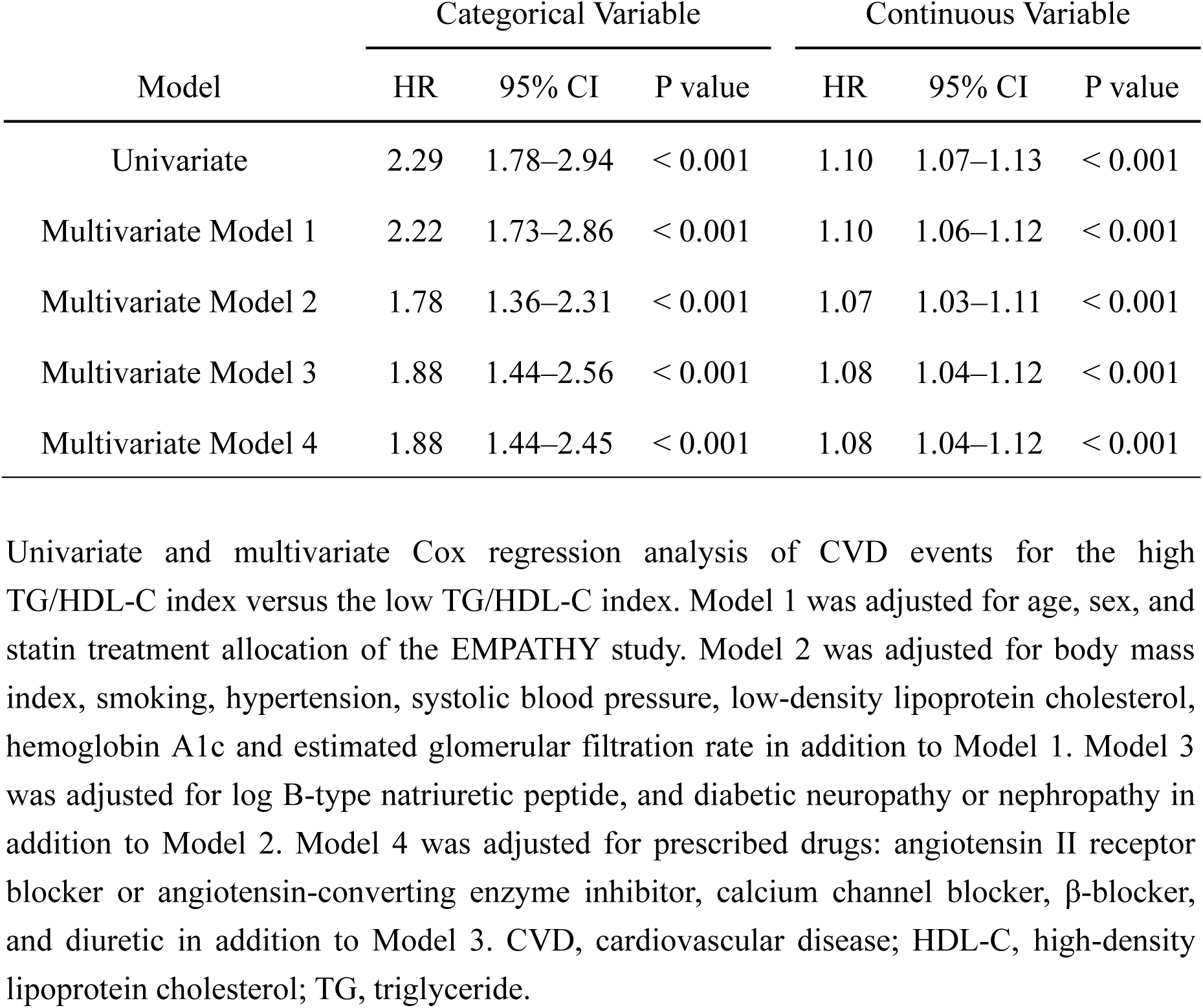
Hazard ratios of CVD events for the high TG/HDL-C index versus the low TG/HDL-C index.

| Model | Categorical Variable |  |  | Continuous Variable |  |  |
| --- | --- | --- | --- | --- | --- | --- |
|  | HR | 95% CI | P value | HR | 95% CI | P value |
| Univariate | 2.29 | 1.78–2.94 | < 0.001 | 1.10 | 1.07–1.13 | < 0.001 |
| Multivariate Model 1 | 2.22 | 1.73–2.86 | < 0.001 | 1.10 | 1.06–1.12 | < 0.001 |
| Multivariate Model 2 | 1.78 | 1.36–2.31 | < 0.001 | 1.07 | 1.03–1.11 | < 0.001 |
| Multivariate Model 3 | 1.88 | 1.44–2.56 | < 0.001 | 1.08 | 1.04–1.12 | < 0.001 |
| Multivariate Model 4 | 1.88 | 1.44–2.45 | < 0.001 | 1.08 | 1.04–1.12 | < 0.001 |

Univariate and multivariate Cox regression analysis of CVD events for the high TG/HDL-C index versus the low TG/HDL-C index. Model 1 was adjusted for age, sex, and statin treatment allocation of the EMPATHY study. Model 2 was adjusted for body mass index, smoking, hypertension, systolic blood pressure, low-density lipoprotein cholesterol, hemoglobin A1c and estimated glomerular filtration rate in addition to Model 1. Model 3 was adjusted for log B-type natriuretic peptide, and diabetic neuropathy or nephropathy in addition to Model 2. Model 4 was adjusted for prescribed drugs: angiotensin II receptor blocker or angiotensin-converting enzyme inhibitor, calcium channel blocker, β-blocker, and diuretic in addition to Model 3. CVD, cardiovascular disease; HDL-C, high-density lipoprotein cholesterol; TG, triglyceride.

To assess whether the TG/HDL index is associated with CVD risk independently of TG and HDL levels, multivariate analyses with additional models were performed. In the multivariate Cox regression analysis adjusted with Model 4 plus TG, TG/HDL-C index ≥2.5 was significantly associated with a high risk of CVD events (HR 1.68, 95% CI 1.24–2.28, P = 0.001), while TG alone was not associated with CVD events **(Table 3)**. Similarly, multivariate analysis adjusted with Model 4 plus HDL-C showed the association of TG/HDL-C index ≥2.5, but not HDL-C, with a high risk of CVD events (HR 1.88, 95% CI 1.38–2.57, P < 0.001). Furthermore, when both TG and HDL-C were added to Model 4, the association of TG/HDL-C index ≥2.5 with a high risk of CVD events was evident (HR 1.67, 95% CI 1.19–2.36, P = 0.003), whereas neither TG nor HDL-C individually showed such an association.

**Table 3.**
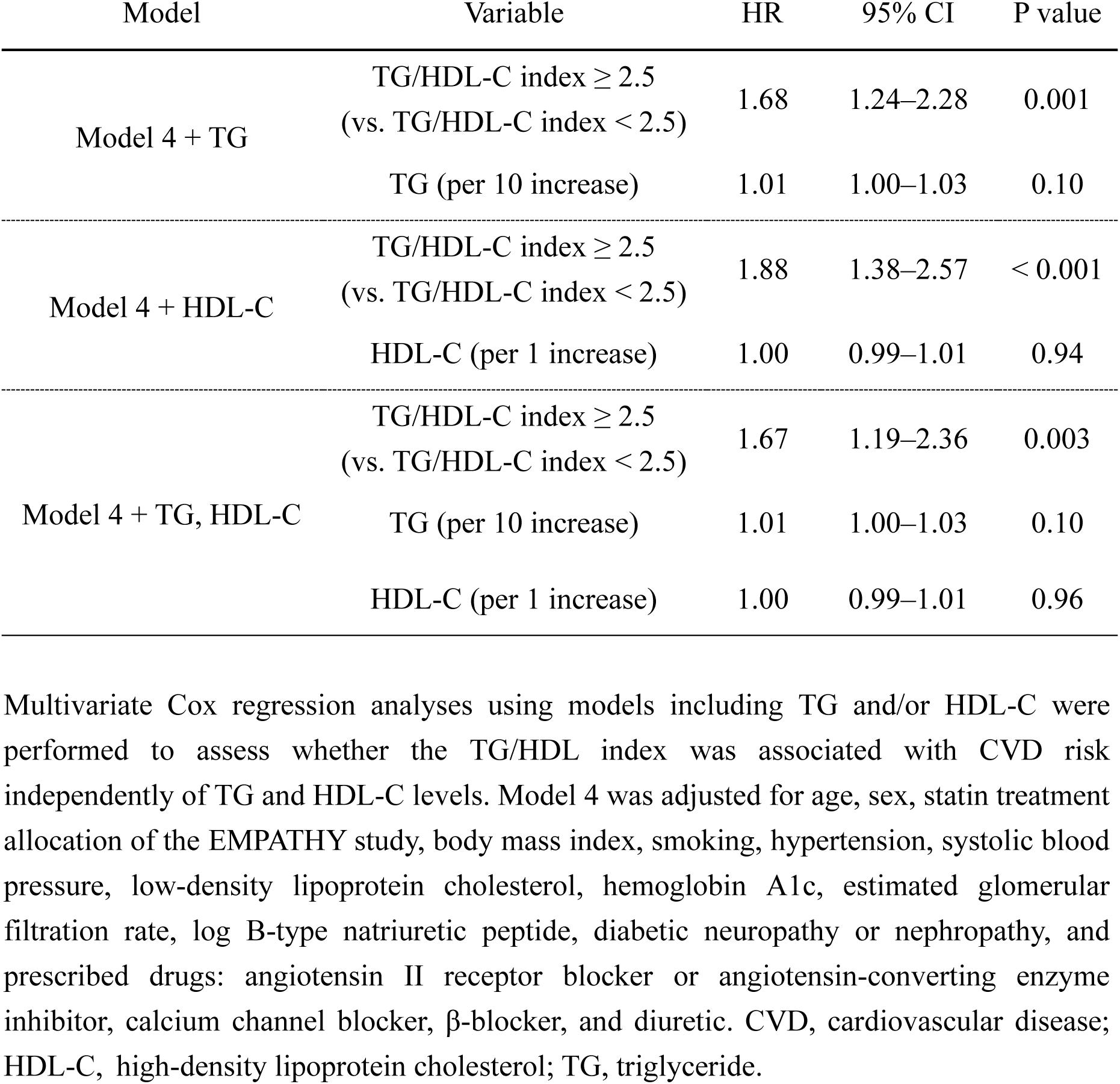
Hazard ratios of CVD events by multivariate Cox regression analyses using models including TG and/or HDL-C.

| Model | Variable | HR | 95% CI | P value |
| --- | --- | --- | --- | --- |
| Model 4 + TG | TG/HDL-C index $\geq 2.5$<br>(vs. TG/HDL-C index $< 2.5$) | 1.68 | 1.24–2.28 | 0.001 |
|  | TG (per 10 increase) | 1.01 | 1.00–1.03 | 0.10 |
| Model 4 + HDL-C | TG/HDL-C index $\geq 2.5$<br>(vs. TG/HDL-C index $< 2.5$) | 1.88 | 1.38–2.57 | $< 0.001$ |
|  | HDL-C (per 1 increase) | 1.00 | 0.99–1.01 | 0.94 |
| Model 4 + TG, HDL-C | TG/HDL-C index $\geq 2.5$<br>(vs. TG/HDL-C index $< 2.5$) | 1.67 | 1.19–2.36 | 0.003 |
|  | TG (per 10 increase) | 1.01 | 1.00–1.03 | 0.10 |
|  | HDL-C (per 1 increase) | 1.00 | 0.99–1.01 | 0.96 |

Multivariate Cox regression analyses using models including TG and/or HDL-C were performed to assess whether the TG/HDL index was associated with CVD risk independently of TG and HDL-C levels. Model 4 was adjusted for age, sex, statin treatment allocation of the EMPATHY study, body mass index, smoking, hypertension, systolic blood pressure, low-density lipoprotein cholesterol, hemoglobin A1c, estimated glomerular filtration rate, log B-type natriuretic peptide, diabetic neuropathy or nephropathy, and prescribed drugs: angiotensin II receptor blocker or angiotensin-converting enzyme inhibitor, calcium channel blocker, β-blocker, and diuretic. CVD, cardiovascular disease; HDL-C, high-density lipoprotein cholesterol; TG, triglyceride.

In the EMPATHY study, CVD events included renal events. Therefore, we also analyzed the association of the TG/HDL-C index with renal events and non-renal CVD events. The TG/HDL-C index ≥2.5 was associated with a high risk of non-renal CVD events (HR 1.99, 95% CI 1.45–2.74, P < 0.001), but not of renal events (HR 1.53, 95% CI 0.96–2.46, P = 0.076) after adjustment using Model 4 **(Table 4)**.

**Table 4.** Hazard ratios of renal events and non-renal CVD events for the high TG/HDL-C index versus the low TG/HDL-C index.

| Outcome | Group | Number of events | Categorical Variable |  |  | Continuous Variable |  |  |
| --- | --- | --- | --- | --- | --- | --- | --- | --- |
|  |  |  | HR | 95% CI | *p* value | HR | 95% CI | *p* value |
| Renal events | TG/HDL index <2.5 | 31/2652 | Reference |  |  | Reference |  |  |
|  | TG/HDL index ≥2.5 | 60/2013 | 1.53 | 0.96–2.46 | 0.076 | 1.04 | 0.97–1.11 | 0.26 |
| Non-renal CVD events | TG/HDL index <2.5 | 69/2652 | Reference |  |  | Reference |  |  |
|  | TG/HDL index ≥2.5 | 106/2013 | 1.99 | 1.45–2.74 | <0.001 | 1.09 | 1.05–1.14 | <0.001 |

Multivariate Cox regression analysis of renal events and non-renal CVD events for the high TG/HDL-C index versus the low TG/HDL-C index using Model 4. Model 4 was adjusted for age, sex, statin treatment allocation of the EMPATHY study, body mass index, smoking, hypertension, systolic blood pressure, low-density lipoprotein cholesterol, hemoglobin A1c, estimated glomerular filtration rate, log B-type natriuretic peptide, diabetic neuropathy or nephropathy, and prescribed drugs: angiotensin II receptor blocker or angiotensin-converting enzyme inhibitor, calcium channel blocker, β-blocker, and diuretic. CVD, cardiovascular disease; HDL-C, high density lipoprotein cholesterol; TG, triglyceride.

### Subgroup analysis

The subgroup analysis based on sex, age (≥65 vs <65 years), BMI (≥25 vs <25 kg/m^2^), HbA1c (≥7.0 vs <7.0%), eGFR (≥60 vs <60 mL/min/1.73 m^2^), TG (≥118 vs <118 mg/dL, based on the median value), HDL-C (≥54 vs <54 mg/dL, based on the median value) and EMPATHY treatment allocation found no significant interaction or trend for interaction between high/low TG/HDL-C index and these factors for the association with the risk of CVD events **(Figure 2)**. However, subgroup analysis based on EMPATHY treatment allocation (intensive or standard statin treatment) showed a trend toward interaction between high/low TG/HDL-C index and EMPATHY treatment allocation for the association with the risk of CVD events (P for interaction = 0.062). Therefore, we performed an additional exploratory analysis to evaluate the effectiveness of intensive statin treatment in the patients stratified by the TG/HDL-C index, the results of which are shown in the next paragraph; while the original EMPATHY study showed that intensive statin treatment did not significantly reduce CVD events in the overall study population.^15^

**Figure 2.**
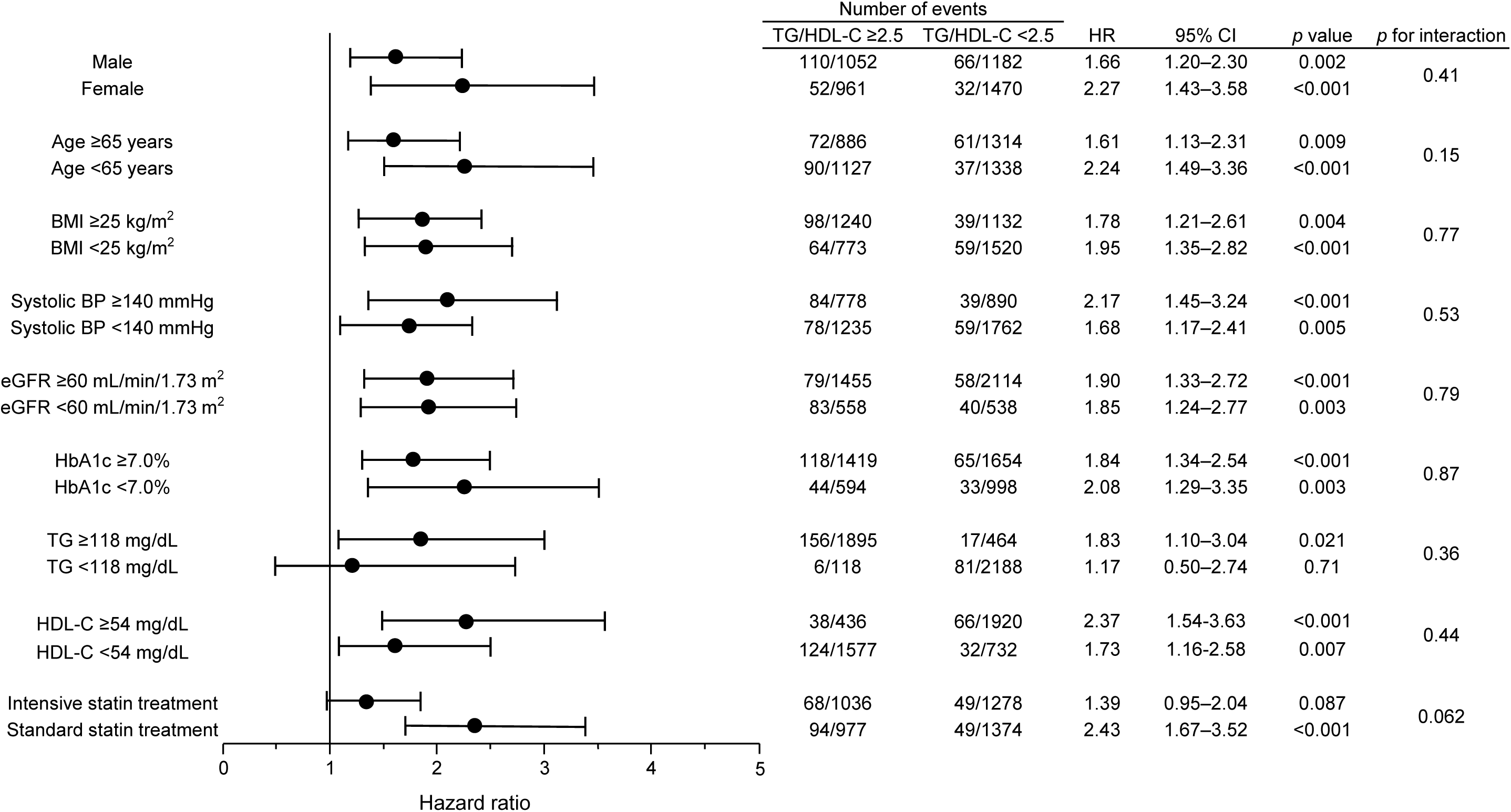
Risk for CVD events in different subgroups. Association between the high TG/HDL-C index and the risk for CVD events in the subgroups stratified by sex, age, BMI, systolic BP, eGFR, HbA1c, TG, HDL-C, and statin treatment allocation in the EMPATHY study. Hazard ratios were calculated by a multivariate Cox proportional hazards model with Model 4. BMI, body mass index; BP, blood pressure; CVD, cardiovascular disease; eGFR, estimated glomerular filtration rate; HbA1c, hemoglobin A1c; HDL-C, high-density lipoprotein cholesterol; TG, triglyceride.

### Different effectiveness of intensive statin treatment based on TG/HDL-C index

**Figure 3** shows the Kaplan-Meier curves comparing the cumulative incidence of CVD events between patients receiving intensive and standard statin treatment in the high TG/HDL-C index group and the low TG/HDL-C index group. In the low TG/HDL-C index group, there was no significant difference in the incidence of CVD events between the intensive and standard statin treatment groups. In the high TG/HDL-C index group, however, the incidence of CVD events was significantly lower in the intensive statin treatment group compared to the standard statin treatment group. Multivariate analysis with a model excluding TG/HDL-C index category from Model 4 also showed that intensive statin treatment was significantly associated with a lower risk of CVD events compared to standard statin treatment in the high TG/HDL-C index group (HR 0.67, 95% CI 0.49–0.91, P = 0.012) but not in the low TG/HDL-C index group (HR 1.09, 95% CI 0.73–1.63, P = 0.68).

**Figure 3.**
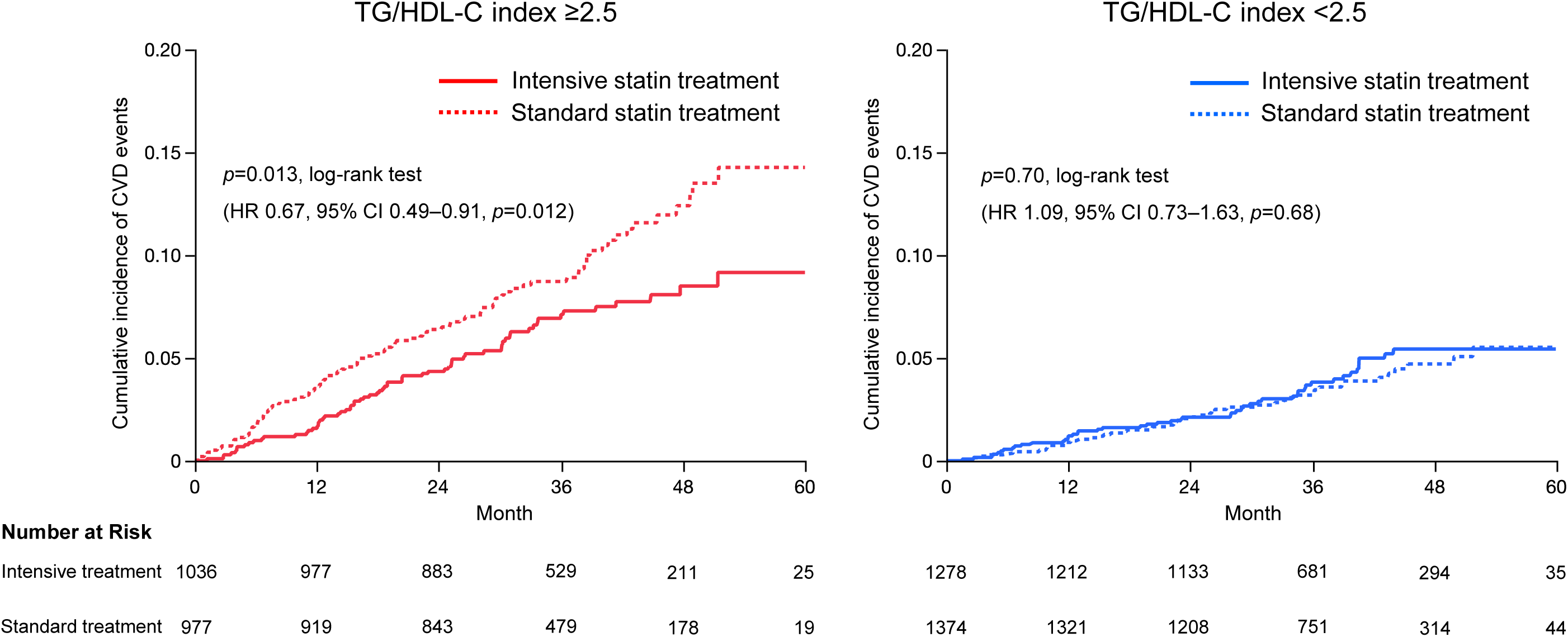
Cumulative incidence of CVD events with intensive and standard statin treatment in the high TG/HDL-C index and low TG-HDL-C index groups. The Kaplan-Meier curves comparing the cumulative incidence of CVD events between the intensive and standard statin treatment groups in the high TG/HDL-C index and low TG/HDL-C index groups. CVD, cardiovascular disease; HDL-C, high-density lipoprotein cholesterol; TG, triglyceride.

## Discussion

This study demonstrates, for the first time, that a high TG/HDL-C index is associated with a high risk of CVD events in patients with T2DM, diabetic retinopathy, and hyperlipidemia without known CVD. In addition to the previous report showing the association of TG/HDL-C index ≥2.5 with a high risk of all-cause mortality and major adverse cardiac events in a limited population with high suspicion of coronary artery disease undergoing coronary angiography,^14^ our findings suggest that TG/HDL-C index ≥2.5 may serve as a predictor of first CVD events in the setting of primary prevention in a larger high-risk population: T2DM patients with diabetic complications and hyperlipidemia. Furthermore, in this study, we investigated the prognostic significance of the TG/HDL-C index considering the levels of TG and HDL-C. Previous studies, including a subanalysis of the EMPATHY study, have shown that TG level predicts the risk of CVD events in patients with T2DM.^16,17^ Even after adjustment for TG and HDL-C, a high TG/HDL-C index was significantly associated with the risk of CVD events in this study. Therefore, the TG/HDL-C index may be a useful predictor independently of TG or HDL-C levels. While patients with T2DM comorbid with diabetic retinopathy are considered as ‘very high risk’, the TG/HDL-C index may be useful for further risk stratification. In addition, our analysis revealed that the effectiveness of intensive statin treatment differed between the high and low TG/HDL-C index groups in the T2DM population. The original EMPATHY trial failed to demonstrate a reduction in the risk of CVD events with intensive statin treatment targeting LDL-C <70 mg/dL compared to standard statin treatment targeting LDL-C ≥100 to <120 mg/dL in patients with T2DM, diabetic retinopathy, and hyperlipidemia in the setting of primary prevention of CVD.^15^ The results of our analysis may present a stratification using the TG/HDL-C index to distinguish patients who may potentially benefit from intensive statin therapy among T2DM patients with diabetic microvascular complications and hyperlipidemia.

The high TG/HDL-C index group was slightly but significantly younger, had a higher proportion of males and current smokers, and had higher BMI, higher blood pressure, lower eGFR, higher TG, lower HDL-C, and lower BNP. Since all of these characteristics, except younger age and lower BNP, are considered risk factors for CVD, the association between the high TG/HDL-C index and higher incidence of CVD events may be partially attributable to these risk factors. However, after adjustment for these potential confounders, the TG/HDL-C index remained significantly associated with a high risk of CVD events. Therefore, a high TG/HDL-C index itself may be an independent predictor of first CVD in the high-risk T2DM population.

There are several possible pathophysiological associations between a high TG/HDL-C index and a high risk of CVD. First, a higher TG/HDL-C index has been suggested to be associated with higher remnant lipoprotein,^18^ which is strongly linked to atherosclerosis.^19,20^ Remnant lipoprotein plays atherogenic, pro-inflammatory, and thrombogenic roles in the development of atherosclerosis leading to CVD: remnant lipoprotein can invade the arterial intima and be absorbed directly by macrophages, promoting foam cell formation and accelerating the progression of atherosclerosis;^21^ remnant lipoprotein is associated with low-grade inflammation as indicated by C-reactive protein;^22^ and remnant lipoprotein promotes the expression of plasminogen activator inhibitor-1 and induces platelet aggregation, increasing the risk of thrombus.^23,24^ Second, a higher TG/HDL-C index is associated with insulin resistance. The TG/HDL-C index is well recognized as a marker of insulin resistance in patients with T2DM.^25–27^ High insulin resistance causes an imbalance in glucose metabolism leading to chronic hyperglycemia and induces oxidative stress, inflammatory responses, and cellular damage, which may also contribute to the development of atherosclerosis and CVD.^28^ Third, high TG and low HDL-C are associated with an increase small dense LDL-C, which is a sub-fraction of LDL-C, but it has a low affinity for LDL-C receptors and its metabolism is delayed. Small dense LDL-C is susceptible to degeneration and, as a result, is not metabolized by the normal metabolic system and supplies cholesterol esters to atherosclerotic lesions. Therefore, a higher TG/HDL-C index may be a condition that facilitates the supply of cholesterol esters to atherosclerotic lesions, thereby promoting arteriosclerosis.^29^

Our analysis showed that intensive statin treatment was associated with a lower risk of CVD compared to standard statin treatment in the high TG/HDL-C index group, but not in the low TG/HDL-C index group **(Figure 3)**. In addition to lowering LDL-C, statins have pleiotropic effects such as suppressing inflammation and improving endothelial function, thereby protecting the vasculature.^30, 31^ As described above, the patients in the high TG/HDL-C index group may have more advanced pathophysiological conditions related to atherosclerosis and CVD; therefore, standard statin treatment may be insufficient to prevent CVD in these patients. In patients with T2DM without known CVD, similar to previous studies showing that the effectiveness of intensive statin treatment was not equivalent when patients were stratified based on blood pressure or visit-to-visit blood pressure variability,^32,33^ the present findings suggest that high TG/HDL-C may be an indicator of patients who will benefit from intensive statin therapy. In this study, the TG/HDL-C index was calculated using baseline TG and HDL-C levels before the statin treatment allocation, which were measured after 4–8 weeks of the run-in statin treatment targeting LDL-C ≥100 to <120 mg/dL according to the protocol of the EMPATHY trial. Therefore, even after statin treatment, i.e., after lowering LDL-C levels, the TG/HDL-C index may reflect a potential risk for CVD events and identify patients for intensive statin treatment in high-risk T2DM patients.

Considering the reduction in the risk of CVD events observed in the high TG/HDL-C index group with intensive statin treatment in our study, and the findings from previous studies investigating lipid-lowering therapy targeting TG, the TG/HDL-C index might be useful in guiding strategies for lipid-lowering therapy. While fenofibrate has shown limited efficacy when used solely in the setting of hypertriglyceridemia,^6–8^ it has demonstrated efficacy when hypertriglyceridemia and hypo-HDL cholesterolemia coexist under statin therapy.^7,34,35^ Similarly, eicosapentaenoic acid (EPA) did not show a primary preventive effect in a clinical study of Japanese subjects,^36^ but demonstrated efficacy in a population with both hypertriglyceridemia and hypo-HDL cholesterolemia.^37^ Furthermore, primary preventive effects of EPA against CVD have been observed in patients with hyper-non-HDL-cholesterol and in patients with both impaired glucose metabolism and hyperglycemia.^38^ These studies may suggest that not only intensive statin treatment but also treatment aimed at lowering TG may be effective in patients with hyper-non-HDL cholesterolemia and concomitant hypo-HDL cholesterolemia, i.e., those with a high TG/HDL-C index.

Although this study analyzed the dataset of the EMPATHY study, which was a prospective, randomized-controlled study including approximately 5000 patients with T2DM and diabetic retinopathy without a history of CVD, we acknowledge some limitations in this study. First, this study was a post-hoc study, which may introduce potential biases or limitations inherent to retrospective studies. Second, other populations, such as patients with T2DM and diabetic nephropathy and/or neuropathy but without diabetic retinopathy, were not included in our analysis. However, given that the risk of cardiovascular events is generally considered to be similar among T2DM patients with each diabetic microvascular complication,^39^ the results of this analysis may applicable to T2DM patients with diabetic complication(s) other than retinopathy. Third, all patients in this study were prescribed statins at baseline and during the follow-up period. Therefore, the association between the TG/HDL-C index and the risk of CVD events may differ in other populations, such as patients not taking statins. Further studies are warranted to elucidate the generalizability and usefulness of the TG/HDL-C index in assessing CVD risk and guiding lipid-lowering therapy strategy.

## Conclusions

A TG/HDL-C index ≥2.5 was associated with a high risk of composite CVD events, renal events, and non-renal CVD events in patients with T2DM and diabetic retinopathy without a history of CVD. Intensive statin treatment was associated with a lower risk of CVD events compared to standard statin treatment in the high TG/HDL-C index group, but not in the low TG/HDL-C index group. These findings suggest that the TG/HDL-C index may be a novel marker for the prediction of CVD events and an indicator of patients who may benefit from intensive statin treatment.

## Data Availability

The datasets generated during and/or analysed during the current study are not publicly available due to ethical restrictions but are available from the corresponding author on reasonable request.

## Clinical Perspective

### What Is New?

- Among 4665 high-risk diabetic patients with retinopathy and hyperlipidemia without known cardiovascular disease (CVD), a high triglyceride/high-density lipoprotein cholesterol (TG/HDL-C) index (≥2.5) was significantly associated with increased CVD risk compared to a low TG/HDL-C index (<2.5) in a dataset from the EMPATHY trial, which compared intensive statin treatment with standard treatment in this population.
- A subgroup analysis showed a trend toward interaction between the TG/HDL-C index and EMPATHY treatment allocation for CVD risk, with an exploratory analysis revealing that intensive statin treatment was associated with reduced CVD risk compared to standard treatment in the high TG/HDL-C index group, but not in the low TG/HDL-C index group.

### What Are the Clinical Implications?

- The TG/HDL-C index may be a novel predictor of CVD and an indicator of patients who may benefit from intensive statin treatment in diabetic patients.

## Non-standard Abbreviations and Acronyms

ACEI: angiotensin converting enzyme inhibitor
ARB: angiotensin II receptor blocker
BMI: body mass index
BP: blood pressure
CCB: calcium channel blocker
CVD: cardiovascular disease
eGFR: estimated glomerular filtration rate
HbA1c: hemoglobin A1c
HDL-C: high density lipoprotein cholesterol
LDL-C: low density lipoprotein cholesterol
T2DM: type 2 diabetes mellitus
TG: triglyceride

## Acknowledgements

None.

## Funding

This research received no specific grant from any funding agency in the public, commercial, or not-for-profit sectors.

## Conflicts of interest

K.S. reports grants from Daiichi Sankyo, Nippon Boehringer Ingelheim, and Otsuka Medical Devices. H.I. reports personal fees from SBI Pharmaceuticals, Wakunaga Pharmaceutical, NIPRO, Meiji, Taisho Pharmaceutical, Ono Pharmaceutical, Kowa, Takeda Pharmaceutical, Daiichi Sankyo, and Novartis Pharma. I.K. reports grant and/or personal fees from Idorsia Pharmaceuticals Japan, Daiichi Sankyo, Takeda Pharmaceutical, Mitsubishi Tanabe Pharma, Teijin Pharma, AstraZeneca, Kowa, MSD, Otsuka Pharmaceutical, Ono Pharmaceutical, Nippon Boehringer Ingelheim, Novartis Pharma, and Bayer Yakuhin. H.T. reports grants and/or personal fees from Daiichi Sankyo, Novartis Pharma, Otsuka Pharmaceutical, Pfizer Japan, Mitsubishi Tanabe Pharma, Teijin Pharma, Nippon Boehringer Ingelheim, AstraZeneca, Ono Pharmaceutical, Kowa, IQVIA Service Japan, MEDINET, Medical Innovation Kyushu, Bayer Yakuhin, Johnson & Johnson, NEC, Nippon Rinsho and Japanese Heart Failure Society. K.A. received grants from Mochida Pharmaceutical and Actelion Pharmaceuticals Japan. Other authors report no conflicts of interest.

## Notes

### Author Declarations

This study was approved by the ethical committee of Kyushu University (No. 2020-445).

